# Sparse Dimensionality Reduction Approaches in Mendelian Randomization with highly correlated exposures

**DOI:** 10.1101/2022.06.15.22276455

**Authors:** Vasilios Karageorgiou, Dipender Gill, Jack Bowden, Verena Zuber

## Abstract

Multivariable Mendelian randomization (MVMR) is an instrumental variable technique that generalizes the MR framework for multiple exposures. Framed as a linear regression problem, it is subject to the pitfall of multi-collinearity. The bias and efficiency of MVMR estimates thus depends on the correlation of exposures. Dimensionality reduction techniques such as principal component analysis (PCA) provide transformations of all the included variables that are effectively uncorrelated. We propose the use of sparse PCA (sPCA) algorithms that create principal components of subsets of the exposures and may provide more interpretable and reliable MR estimates. The approach consists of three steps. We first apply a sparse dimension reduction method and transform the variant-exposure summary statistics to principal components. We then choose a subset of the principal components based on data-driven cutoffs, and estimate their strength as instruments with an adjusted *F*-statistic. Finally, we perform MR with these transformed exposures. This pipeline is demonstrated in a simulation study of highly correlated exposures and an applied example using summary data from a genome-wide association study of 118 highly correlated lipid metabolites. As a positive control, we tested the causal associations of the transformed exposures on CHD. Compared to the conventional inverse-variance weighted MVMR method and a weak-instrument robust MVMR method (MR GRAPPLE), sparse component analysis achieved a superior balance of sparsity and biologically insightful grouping of the lipid traits.

**Key Messages:**

- In multivariable MR, investigation of multiple highly correlated exposures can hinder the efficiency of the estimators and mask true associations.
- Dimensionality reduction approaches such as principal component analysis (PCA) appear to be effective in summarising the variant-exposure summary statistics data in an example of correlated metabolite data.
- Sparse PCA approaches have the additional benefit of providing interpretable PCs as only a few exposures contribute to each. This benefit is shown in simulation studies where there is a gain in accuracy over PCA.
- In a positive control analysis, the sparse PCs were representing biologically meaningful groups of metabolites (VLDL, LDL, HDL) and in general were associated with coronary heart disease in an anticipated mannert.

## Introduction

Mendelian randomization (MR) uses genetic variants as instrumental variables (IVs) to investigate the causal effect of a genetically predicted exposure on an outcome of interest^1^. The random allocation of genetic variants at conception helps overcome the environmental confounding that can hinder traditional epidemiological study designs based on observational associations. Genetically predicted levels of lifelong exposures are also less likely to be affected by reverse causation, as genetic variants are allocated before onset of the outcomes of interest.

When evidence suggests that multiple correlated phenotypes may contribute to a health outcome, multi-variable MR (MVMR), an extension of the basic univariable approach, can disentangle more complex causal mechanisms and shed light on mediating pathways. An example of this would be investigation into the effect of various lipid traits on coronary heart disease (CHD) risk^2^. While MVMR can model correlated exposures, it performs suboptimally when there are highly correlated exposures due to multi-collinearity. This can be equivalently understood as a problem of conditionally weak instruments^3^. That is, the genetic variants used to proxy all exposures have to be strongly associated with each exposure conditionally on all the other included exposures. An assessment of the extent to which this assumption is satisfied can be made using the conditional F-statistic, with a value of 10 being considered sufficiently strong^3^. In settings when multiple highly correlated exposures are analysed, a set of genetic instruments are much more likely to be conditionally weak instruments. In this event, causal estimates can be subject to extreme bias and are therefore unreliable. Estimation bias can be addressed to a degree by fitting weak-instrument robust MVMR methods^4,5^, but usually at the cost of a further reduction in precision. Furthermore, MVMR models investigate causal effects for each individual exposure, under the assumption that it is possible to intervene and change each one whilst holding the others fixed. In the high dimensional, highly correlated exposure setting, this is potentially an unachievable intervention in practice.

Our aim in this paper is instead to use dimensionality reduction approaches to concisely summarise a set of highly correlated genetically predicted exposures into a smaller set of independent principal components (PCs). We then perform MR directly on the PCs, thereby estimating their effect on health outcomes of interest. We additionally suggest employing sparsity methods to reduce the number of exposures that contribute to each principal component, in order to improve their interpretability in the resulting factors.

Using summary genetic data for multiple hightly correlated lipid fractions and CHD^6,7^, we first illustrate the pitfalls encountered by the standard MVMR approach. We then apply a range of sparse PCA methods within an MVMR framework to the data. Finally, we examine the comparative performance of the sparse PCA approaches in a detailed simulation study, in a bid to understand which ones perform best in this setting.

## Methods

### Workflow Overview

Our proposed analysis strategy is presented in Figure 1. Using summary statistics for the singlenucleotide polymorphism (SNP)-exposure (*γ*) and SNP-outcome (Γ) associations, where *γ* exhibits strong correlation, we initially perform a principle component analysis (PCA) on *γ*. Additionally, we perform multiple sparse PCA modalities that aim to provide sparse loadings that are more interpretable (block 3, Fig. 1). The choice of the number of principal components (PCs) is guided by permutations or an eigenvalue threshold. Finally, the PCs are used in place of *γ* in an IVW MVMR meta-analysis to obtain an estimate of the causal effect of the PC on the outcome. Similar to PC regression and in line with unsupervised methods, the outcome (SNP-outcome associations (Γ) and corresponding standard error (*SE*_Γ_)) is not transformed by PCA and is used in the second-step MVMR in the original scale. In the real data application and in the simulation study, the best balance of sparsity and statistical power was observed for the method of sparse component analysis (SCA)^8^. This favoured method and the related steps are coded in an *R* function and are available at github (https://github.com/vaskarageorg/SCA_MR/).

**Figure 1.**
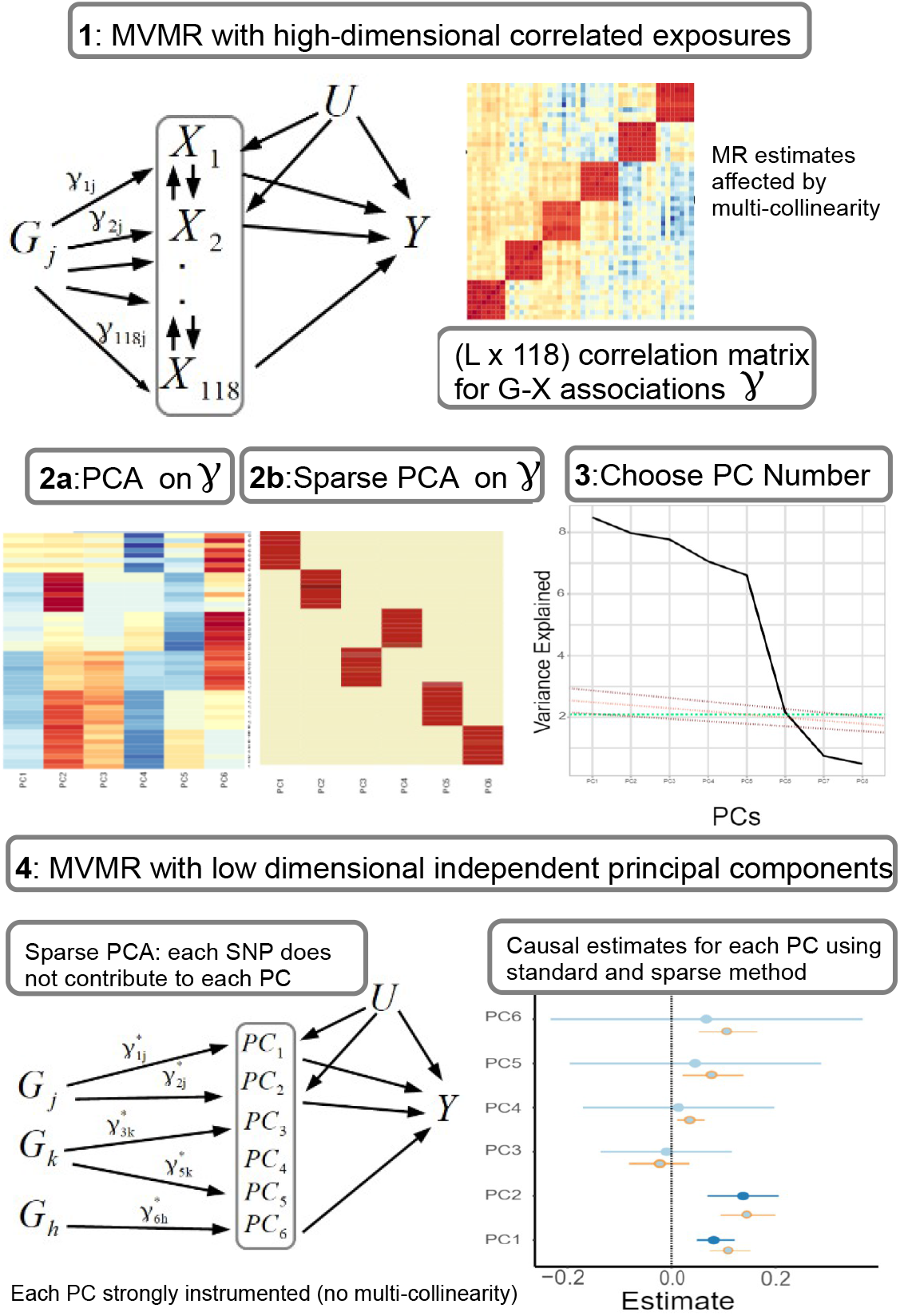
Proposed workflow. In step 3, the yellow colouring represents values close to zero or exactly zero in the right plot for the sparse PCA loadings. In step 4, the choice of the number of PCs that are carried forward for analysis is visualised (green: eigenvalue cutoff^9^; red: permutation-based approach). In step 5, the causal estimates with PC scores (orange) and sparse PC scores (blue) are presented. X: exposures; Y: outcome; k: number of exposures; PCA: principal component analysis; MVMR: multivariable MR

### Data

The risk factor dataset reports the associations of 148 genetic variants (SNPs) with 118 NMR-measured metabolites^6^. This study reports a high-resolution profiling of mainly lipid subfractions. Fourteen size/ density classes of lipoprotein particles (ranging from extra small (XS) high density lipoprotein (HDL) to extra-extra-large (XXL) very low density lipoprotein (VLDL)) were available and, for each class, the traits of total cholesterol, triglycerides, phospholipids, and cholesterol esters content, and the average diameter of the particles were additionally provided. Through the same procedure, estimates from NMR on genetic associations of amino acids, apolipoproteins, fatty and fluid acids, ketone bodies and glycerides were also included. Instrument selection for this dataset has been previously described^10^. Namely, 185 variants were selected, based on association with either one of: LDL-cholesterol, HDL-cholesterol or triglycerides in the external sample of the Global Lipid Genetics Consortium at the genome-wide level *p <* 5 × 10^−8^)^11^. Then, this set was pruned to avoid inclusion of genes in linkage disequilibrium (threshold: *r*^2^ *<* 0.05) and filtered (variants in distance less than 1 megabase pairs were excluded) resulting in the final set. This pre-processing strategy was performed with a view to study CHD risk.

Positive control outcome assessment is recommended in MR as an approach of investigating a risk factor that has an established causal effect on the outcome of interest^12^. We used CHD as a positive control outcome, given that lipid fractions are known to modulate its risk, with VLDL-and LDL-related traits being positively associated with CHD and HDL-related traits negatively^12^. Summary estimates from the CARDIoGRAMplusC4D Consortium and UK Biobank meta-analysis^7,13^ were used. For both datasets, a predominantly European population was included, as otherwise spurious results could have arisen due to population-specific, as opposed to CHD-specific, differences^14^.

### MR Assumptions

The first assumption (IV1) states that IVs should be associated with the exposure. The second assumption (IV2) states that the IVs should be independent of all confounders of the risk factor-outcome association (IV2) and, finally, independent of the outcome conditional on the exposure and the confounders (IV3). The validity of the final assumption cannot be directly tested with the available data. For the inferences to be valid, it is necessary that the three IV assumptions apply^15^. In situations when IV3 is not deemed likely, additional risk factors that are possible mediators can be introduced in a multivariable MR (MVMR) model^2^. Additional assumptions should hold for MVMR results to be valid. Particularly, a) all included exposures have to be associated with at least one of the IVs, and, b) there should be no evidence of multicollinearity. If there is significant overlap (if *G*_1_ and *G*_2_ associate with both the exposures in Fig. 2), this may result in conditionally weak instruments^4^. The latter assumption and the way it limits eligibility of high-dimensional, correlated exposures is a key motivation for the present work.

**Figure 2.**
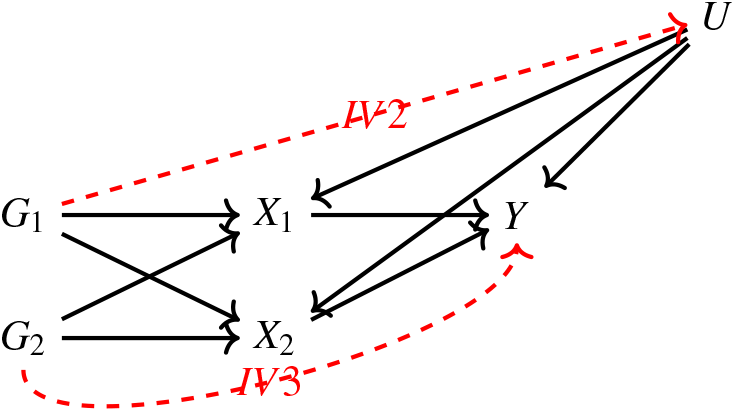
Directed acyclic graph (DAG) for the MVMR assumptions. IV2, IV3: instrumental variable assumptions 2 and 3.

### Univariable (UVMR) & Multivariable MR (MVMR)

To examine how each of the metabolites is causally associated with CHD, a univariable MR (UVMR) approach was first undertaken under the two-sample summary data framework. This uses SNP-exposure and SNP-outcome GWAS summary statistics (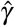 and 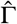 respectively), obtained by regressing the exposure or outcome trait on each SNP individually. Usually, an additional adjustment has been made for variables such as age, gender, and genetic principal components. A UVMR analysis is the most straightforward way to obtain an estimate for the causal effect of the exposure, but is only reliable if all SNPs are valid instruments. However, in the Kettunen dataset, where the exposure set comprises 118 highly correlated lipid fractions, one variant may influence multiple exposures. This will generally invalidate a UMVR analysis and a multivariable MR (MVMR) approach is more appropriate.

Estimating equation (1) provides a general means for representing the mechanics of a UMVR or MVMR analysis:

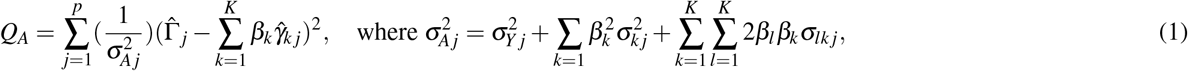

where

- 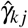 represents the association of SNP *j* with exposure *k*, with variance 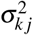;
- 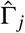 represents the association of SNP *j* with the outcome, with variance 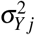;
- *β*_*k*_ represents the causal effect of exposure *k* on the outcome to be estimated.
- *σ*_*lk j*_ represents 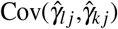.

In a UMVR analysis there is only one exposure, so that *K*=1, whereas in a MVMR analysis *K* ≥ 2 (or in the case of the Kettunen data, *K*=118). In an IVW UMVMR or MVMR analysis, the causal effect parameters estimated are obtained by finding the values 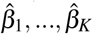 that minimise *Q*_*A*_, under the simplifying assumption that 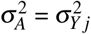. This is justified if all causal effects are zero, or if the uncertainty in the SNP-exposure associations is negligible (the No Measurement Error (NOME) assumption). In the general MVMR context, the NOME assumption is approximately satisfied if the conditional *F*-statistics (CFS) for each exposure are large, but if it is not, then IVW MVMR estimates will suffer from weak instrument bias. For exposure *k, CFS*_*k*_ takes the form

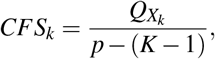

where:

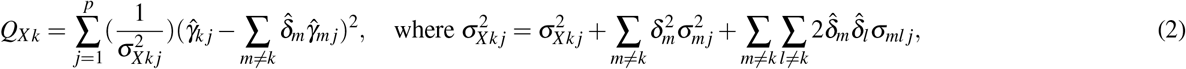

and where the 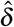 are obtained by OLS regression. CFS statistics will generally be small (indicating weak instrument bias) whenever there is a high degree of correlation between the SNP-exposure association estimates. Such a high correlation is an indication of multi-colinearity. This would be the case in an MVMR analysis of all 118 lipid fractions in the Kettunen data. One option to address this would be to use weak instrument robust MVMR, such as MR-GRAPPLE^5^. This method performs estimation using the full definition of 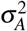 in (1). It can work well for MVMR analyses of relatively small numbers of exposures (e.g. up to 10) and relatively weak instruments (CFS statistics as low as 3), but the dimension and high correlation of the Kettunen data is arguably too challenging. This motivates the use of dimension reduction techniques, such as PCA.

### Dimension reduction via PCA

The PCA method used was a singular value decomposition of the *p* × *K* matrix of SNP-exposure associations 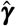 as:

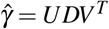

where *U* and *V* are orthogonal matrices and *D* is a square matrix whose diagonal values are the variances explained by each component and all off diagonal values are 0. V is the loadings matrix and serves as an indicator of the contribution of each metabolite to the transformed space of the PCs. The matrix *UD* (PCs/ scores matrix) is used in the second-step IVW regression in place of *γ*. As *V* estimation does not necessarily guarantee sparsity, certain exposures can contribute to multiple components, making the interpretation more complicated. Therefore, we assessed multiple sparse PCA methods that facilitate this.

#### Sparse PCA (Zou et al)

Sparse PCA by Zou et al.^16^ estimates the loadings matrix through an iterative procedure that progressively penalises exposures so that they do not contribute to certain PCs. In principle, this leads to a more clear picture for the consistency of each PC. This is performed as follows

1. Setting a fixed matrix, the following elastic net problem is solved

*ξ*_*j*_ = *argmin*_*ξ*_ (*α*_*j*_ − *ξ*)^*T*^ *γ*^*T*^ *γ*(*α*_*j*_ − *ξ*) + *λ*_1 *j*_∥*ξ* ∥ + *λ* ∥*ξ* ∥^2^, where *j* is the PC.

2. For a fixed Ξ, *γ*^*T*^ *γ*Ξ = *UDV*^*T*^ is estimated and update *A* = *UV*^*T*^

3. Repeat until convergence

As a result of the additional *λ*_1_∥*ξ* ∥ norm, there is sparsity in the loadings matrix and only some of the SNP-exposure associations *γ* contribute to each PC, specifically a particular subset of highly correlated exposures in *γ*.

#### RSPCA

This approach differs in that it employs a robust measure of dispersion (*Q*_*n*_ estimator)^17,18^. As above, an *L*_1_ norm is used to induce sparsity. For optimisation, the Tradeoff Product Optimization (TPO) was maximised. Its advantage is that it does not impose a single *λ* value on all PCs, thus allowing different degrees of sparsity. The maximum number of iterations was set at 100. In our application of a robust measure of dispersion for transforming the Kettunen data, this could result in data such that outlying associations may not distort the resulting transformations. Outliers in this context are defined only in *γ*, in contrast with the median and mode methods^19,20^.

#### *Sparse Fused Principal Component Analysis (SFPCA)*^*21*^

A method that can in theory exploit distinct correlation structures. The goal is to derive a loadings matrix in which highly positively correlated variables are similar in sign and highly negative ones are opposite. Similar magnitudes also tend to be obtained for those variables that are in the same blocks in the correlation matrix. Like the optimisation in Zou et al.^16^, the following criterion is used

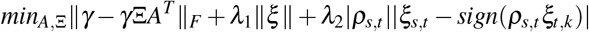

s.t. *A*^*T*^ *A* = *I*_*K*_. The ‘fused’ penalty (last term) purposely penalises discordant loadings for variables that are highly correlated. The choice of the sparsity parameters is based on a BIC criterion.

#### Sparse Component Analysis (SCA)

SCA^8^ is motivated by the relative inadequacy of the classic approaches in promoting significant sparsity. The authors present a solution of the variance maximisation problem in which the eigenvectors are rotated in a way that approximate sparsity is achieved. This is prompted by the fact that the potential loadings for the problem of variance maximisation in sPCA form an entire space if the orthogonality constraint does not hold and, thereby, a loadings solution can be rotated so that it’s (approximately) sparse and explains the same proportion of variance. Then, a soft-thresholding approach is applied on the loadings matrix through constraint with a sparsity parameter. The simulation studies support the increased sparsity in a dataset of gene expression, among other examples^8^.

### Choice of Components

In all dimensionality reduction approaches, there is no upper limit to how many transformed factors can be estimated. However, only some of them are informative in the sense of explaining a meaningful proportion of variance in the original dataset. For this purpose, a permutation-based approach was implemented^22^. The *γ* matrix was randomly reshuffled (permuted) and the sparse PCA method of interest was applied in the permuted set. In this case, the eigenvalue that is computed reflects the eigenvalue obtained under the null of a non-informative component. This process is repeated multiple times (*perm* = 1000) and the mean eigenvalue for all components is stored. Finally, the sparse PCA method is performed in the original *γ* matrix and whichever component has an eigenvalue larger than the mean of the permuted sets is considered informative.

Due to the computational complexity of the permutation method, particularly for SFPCA, an alternate method -the Karlis-Saporta-Spinakis (KS<u>S) c</u>riterion^9^ - was also used. This is based on a simple correction on the minimum non-trivial eigenvalue (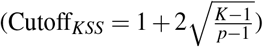). The authors show that the method is not sensitive to non-normal distributions^9^. Although KSS was not compared with the above described permutation approach, it performed better than simpler approaches, such as choosing those PCs whose eigenvalue is larger than 1 (Kaiser criterion), the broken stick method^23^ and the Velicer method^24^.

### Instrument Strength of PCs

In MVMR, the *IV* 1 assumption requires a set of genetic variants that robustly associate with at least one of the exposures *X* (*MR Assumptions*). This is quantified by the *F*-statistic and CFS^4^. As a rule of thumb, if the *F*-statistic surpasses the cutoff of 10, the transformed exposure is strongly instrumented. With summary statistics of the SNP-*X* associations *γ*_*p,k*_ (*p*: SNP, *k*: exposure), the mean *F*−statistic for the exposure *k* is estimated as

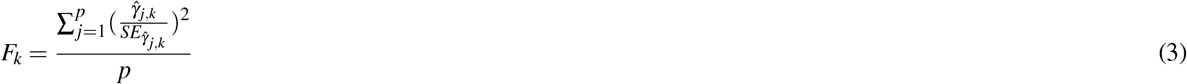

Since we transform *γ* and obtain a matrix of lower dimensionality, formula 3 can’t be used as there is no longer a one-to-one correspondence of the *SE*s with the PCs. Likewise, a conditional *F*−statistic for the PCs also cannot be computed for this reason. We aim to arrive at a modified formula that bypasses this issue. For this purpose, we take advantage of two concepts, first an expression of the *F*−statistic for an exposure *k* (*F*_*k*_) in matrix notation and, second, the use of this expression to estimate *F*−statistics for the PCs (*F*_*PC*_) from *γ* decomposition.

We make the assumption that the uncertainty in the *γ*_*G,XK*_ estimates is similar in all *K* exposures, i.e. *γ*_*G,X*_ uncertainty estimates do not substantially differ among exposures. This is not implausible as the uncertainty is predominantly driven by sample size and minor allele frequency^25^. In specific, the authors of^25^ show that

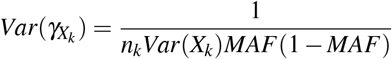

where MAF is the minor allele frequency, *n*_*k*_ is the sample size in exposure *k* and *Var*(*X*_*k*_) is the phenotypic variance. What this means is that, in experiments such as^6^ where *n*_*k*_ is the same across all exposures and *Var*(*X*_*k*_) can be standardised to 1, the main driver of differences in *Var*(*γ*_*XK*_) is differences in MAF. As MAF is the same for each SNP across all exposures, the collation of SEs across exposures per SNP is well motivated.

We can then define a matrix Σ as follows.

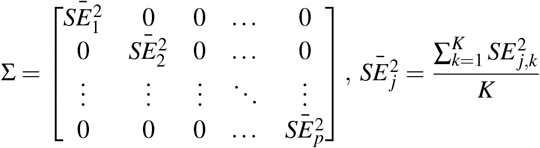

The elements in the diagonal represent the mean variance of *γ* for each SNP and all off-diagonal elements are zero. What is achieved through this is a summary of the uncertainty in the SNP-*X* associations that is not sensitive on the dimensions of the exposures. Instead of Eq. 3, we can then express the vector of the mean *F*-statistics for each exposure *F*_1−*K*_ = [*F*_1_, *F*_2_, …, *F*_*K*_] as

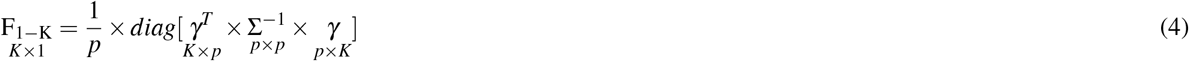

where *γ* is the *matrix* of the SNP-exposure associations. In a simulation study, we generate data under the mechanism in Figure 3a. The strength of association is different in the three exposures. It is observed that the estimates with both methods (Eq. 4 and Eq. 3) align well (Figure 3b), supporting the equivalence of the two formulae.

**Figure 3.**
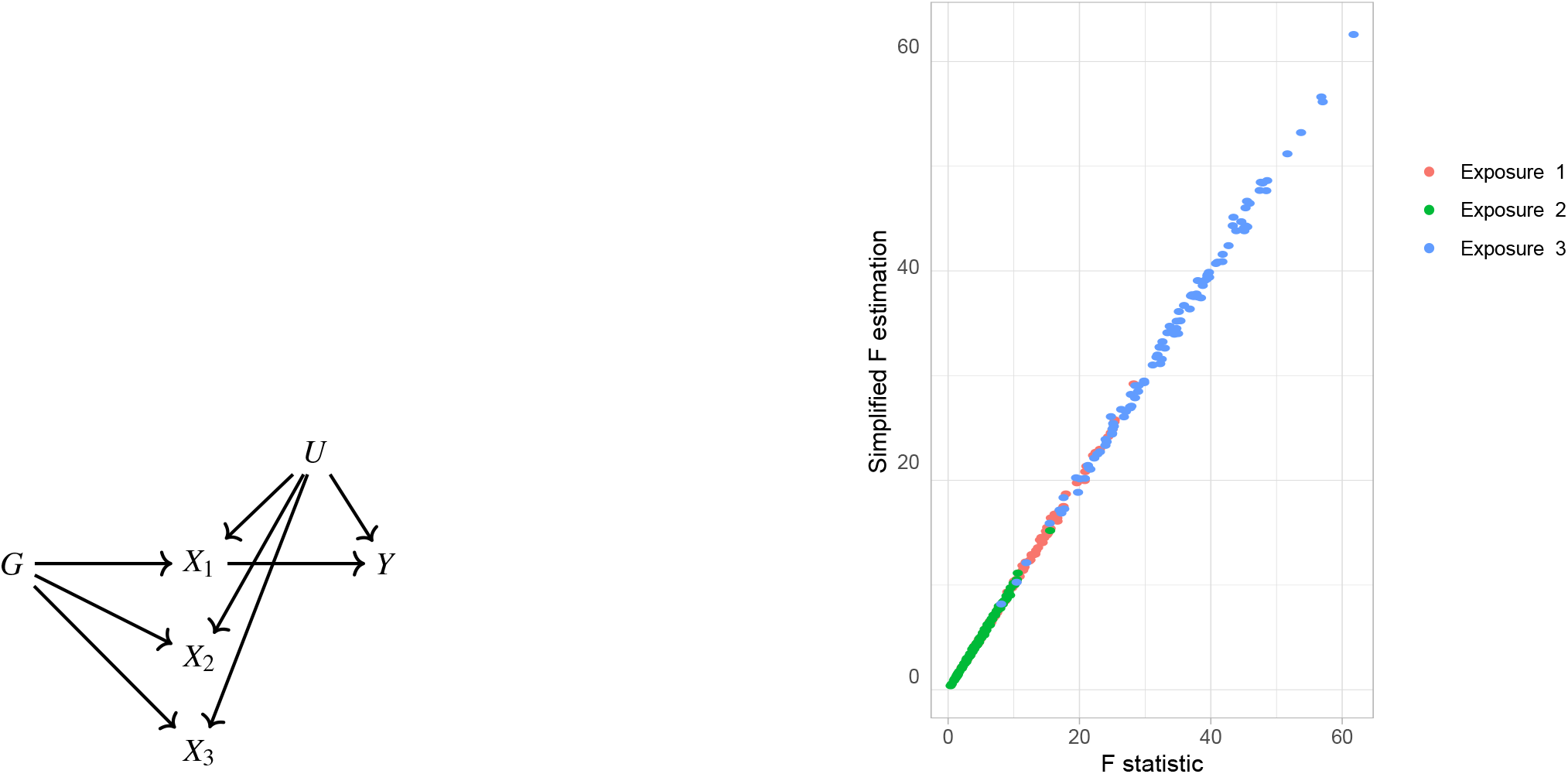
a. Data generating mechanism. Three exposures with different degrees of strength of association with *G* are generated *γ*_1_ = 1, *γ*_2_ = 0.5, *γ*_3_ = 0.1. b. *F*-statistic for the three exposures *X*_1_, *X*_2_, *X*_3_ as estimated by the formulae in Eq. 4 (horizontal axis) and Eq. 3 (vertical axis).

Our second aim is to use this matrix notation formula for the *F*−statistic to quantify the instrument strength of each PC with the respective *F*−statistic (*F*_*PC*_). As presented above, we are not limited by the dimensions of point estimates and uncertainty matching exactly and we can use the formula in Eq. 4 and substitute ***γ*** with the PCs. For the PCA approach, where we decompose ***γ*** as ***γ*** = *UDV*^*T*^ and carry forward *M << K* non-trivial PCs, we use the matrix *UD* in place of ***γ***. Then, the mean *F*_*PC*_ can be estimated as follows.

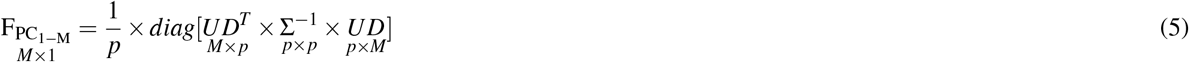

The vector 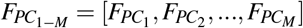 contains the *F*_*PC*_ statistics for the *M* PCs. In a similar manner, we estimate *F*_*PC*_ for the sparse PCA methods but, instead of the scores matrix *UD*, we use the scores of the sparse methods. We illustrate the performance of this approach in a simulation study with an identical configuration for exposure generation as the one presented in Figure 8. In a configuration with *b* = 6 blocks of *p* = 30 genetically correlated exposures (Figure 7), we vary the strength of association *γ* per block. This way, the first block has the highest strength of association and the last block the lowest, quantified by a lower mean *F*−statistic in the exposures of this block (red, Figure 4). The instrument strength of the PCs and the sPCs follow closely the corresponding *F*−statistics of the individual exposures; in other word, in a PC of five exposures, 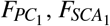 and *F*_1−5_ align well (Figure 4).

**Figure 4.**
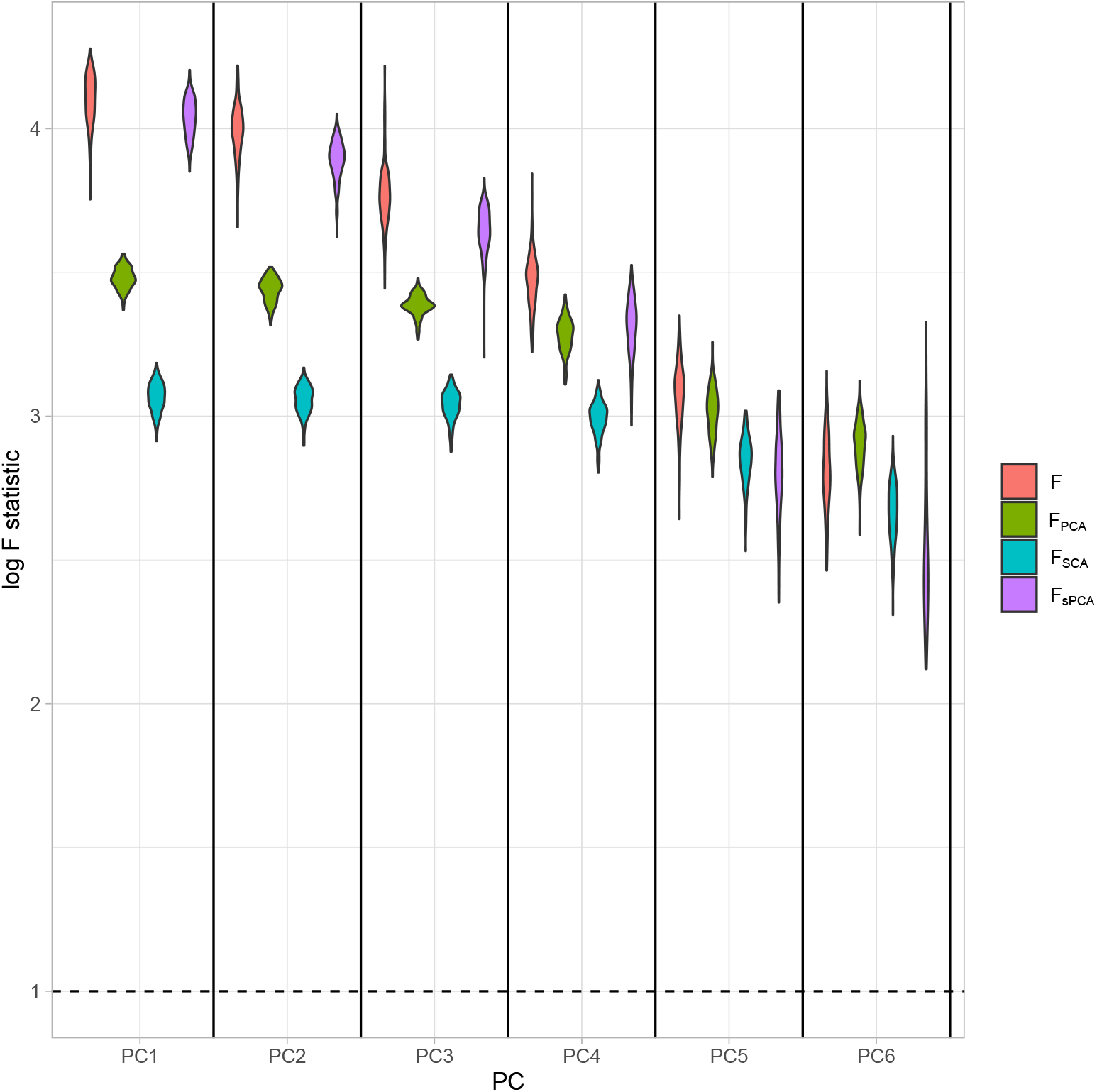
Distributions of the *F*−statistics in PCA methods and individual (not transformed) exposures. Exposure data in different blocks are simulated with a decreasing strength of association and the correlated blocks map to PCs. Each distribution represents the *F*−statistics for each PC. In the case of the individual exposures (red), the distributions represent the *F*−statistics for the corresponding exposures. Individual: individual exposures without any transformation; PCA: *F*−statistics for PCA; SCA: sparse component analysis^8^; sPCA: sparse PCA as described by Zou et al.^16^

**Figure 5.**
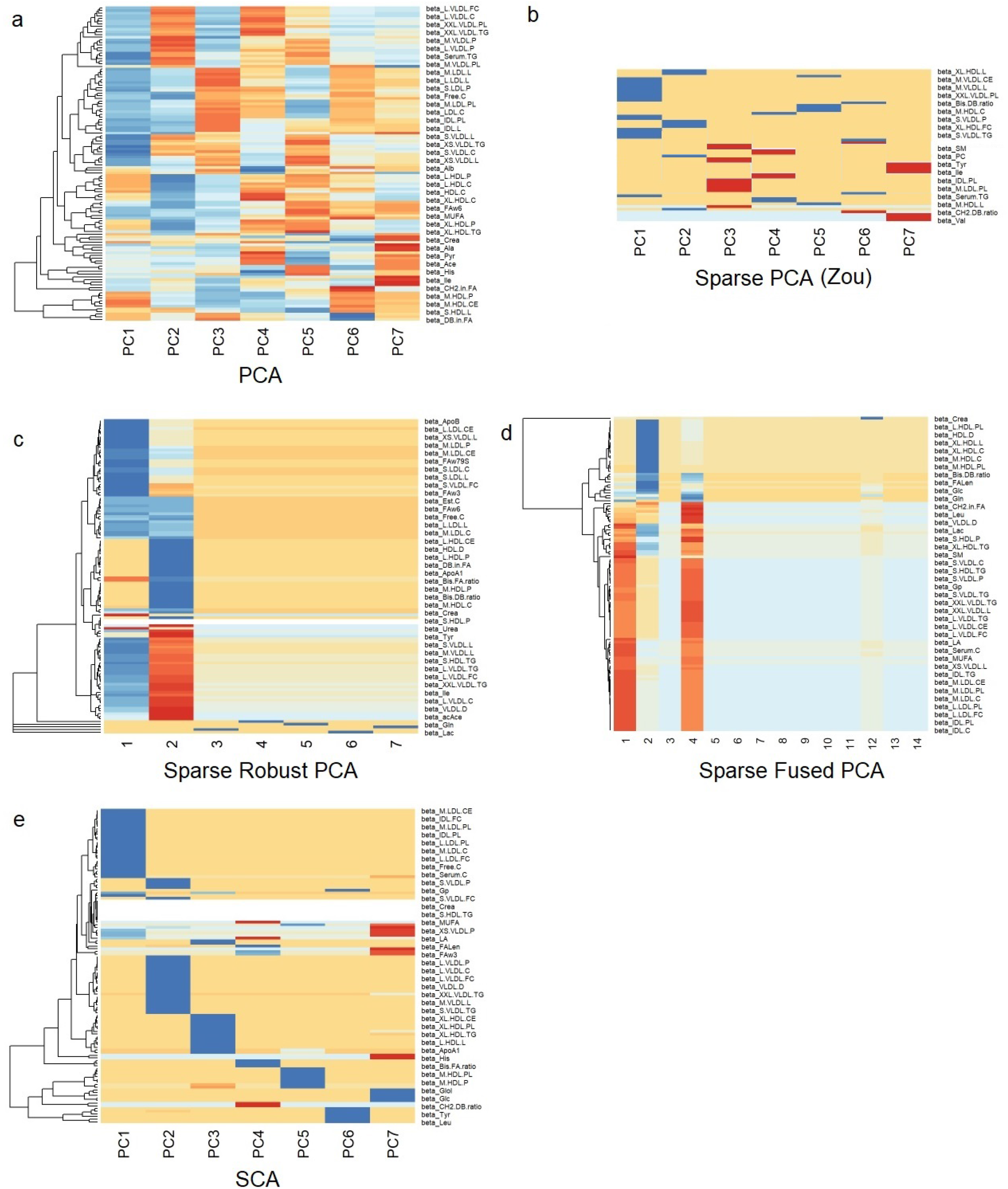
Heatmaps for the loadings matrices in the Kettunen dataset for all methods (one with no sparsity constraints (a), four with sparsity constraints under different assumptions (b-e)). Blue: positive loading; Red: negative loading; Yellow: zero.

**Figure 6.**
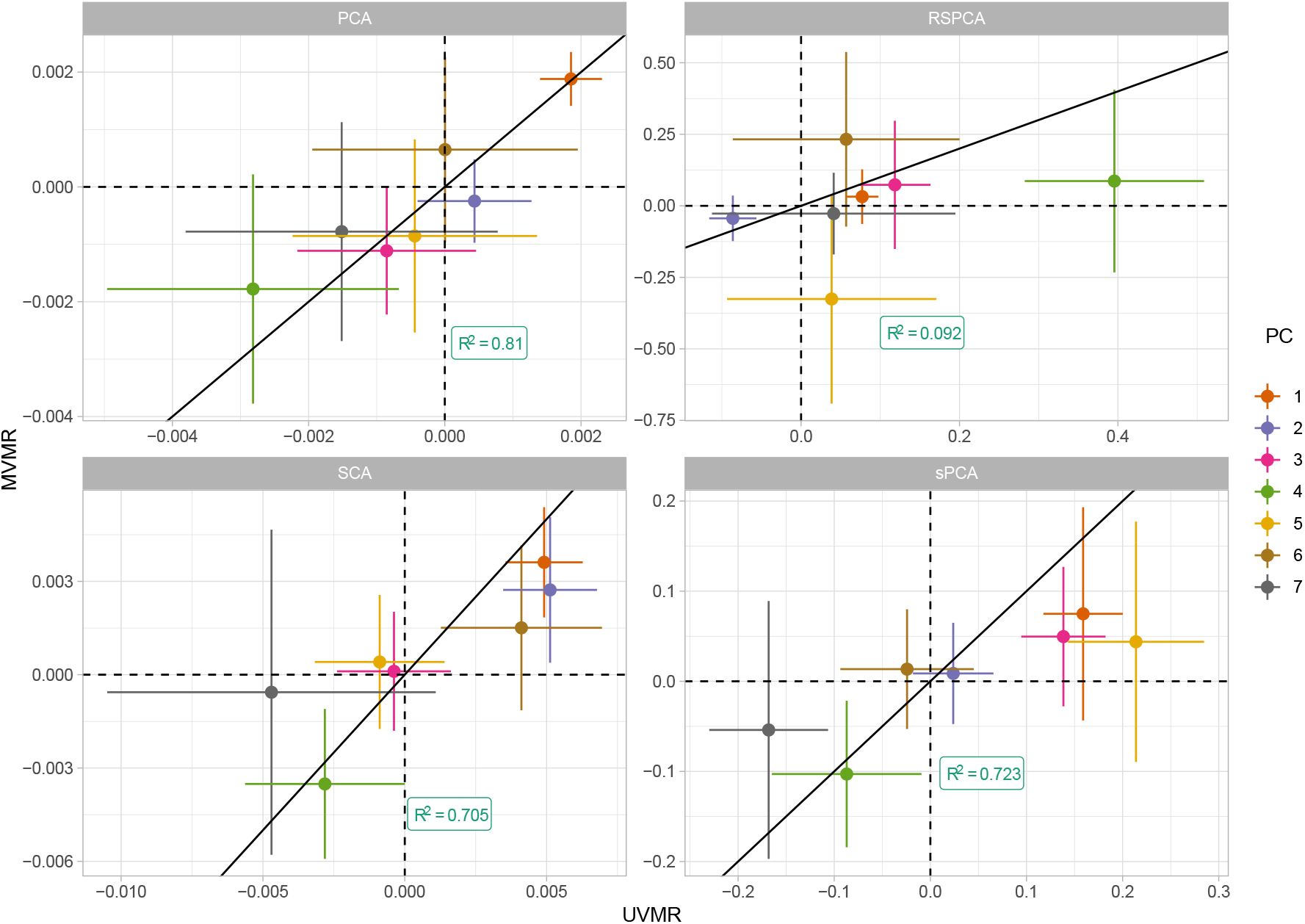
Extrapolated ROC curves for all methods. SCA: Sparse Component Analysis^8^; sPCA: sparse PCA (Zou et al.)^16^; RSPCA: robust sparse PCA^26^; PCA: principal component analysis; MVMR: multivariable MR; MVMR_B: MVMR with Bonferroni correction.

**Figure 7.**
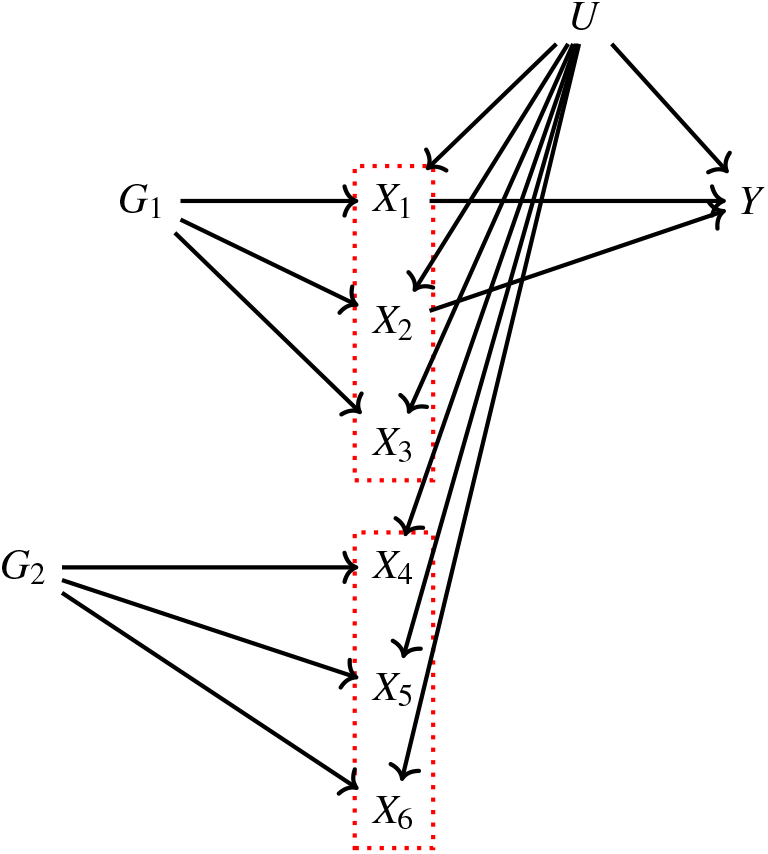
Data generating mechanism for the simulation study. For the sake of clarity, only six exposures are presented. In red boxes, the exposures that are correlated due to a shared genetic component are highlighted. The aim of dimension reduction is to collate *X*_1_, *X*_2_, *X*_3_ to a single component and *X*_4_, *X*_5_, *X*_6_ to a second independent component.

**Figure 8.**
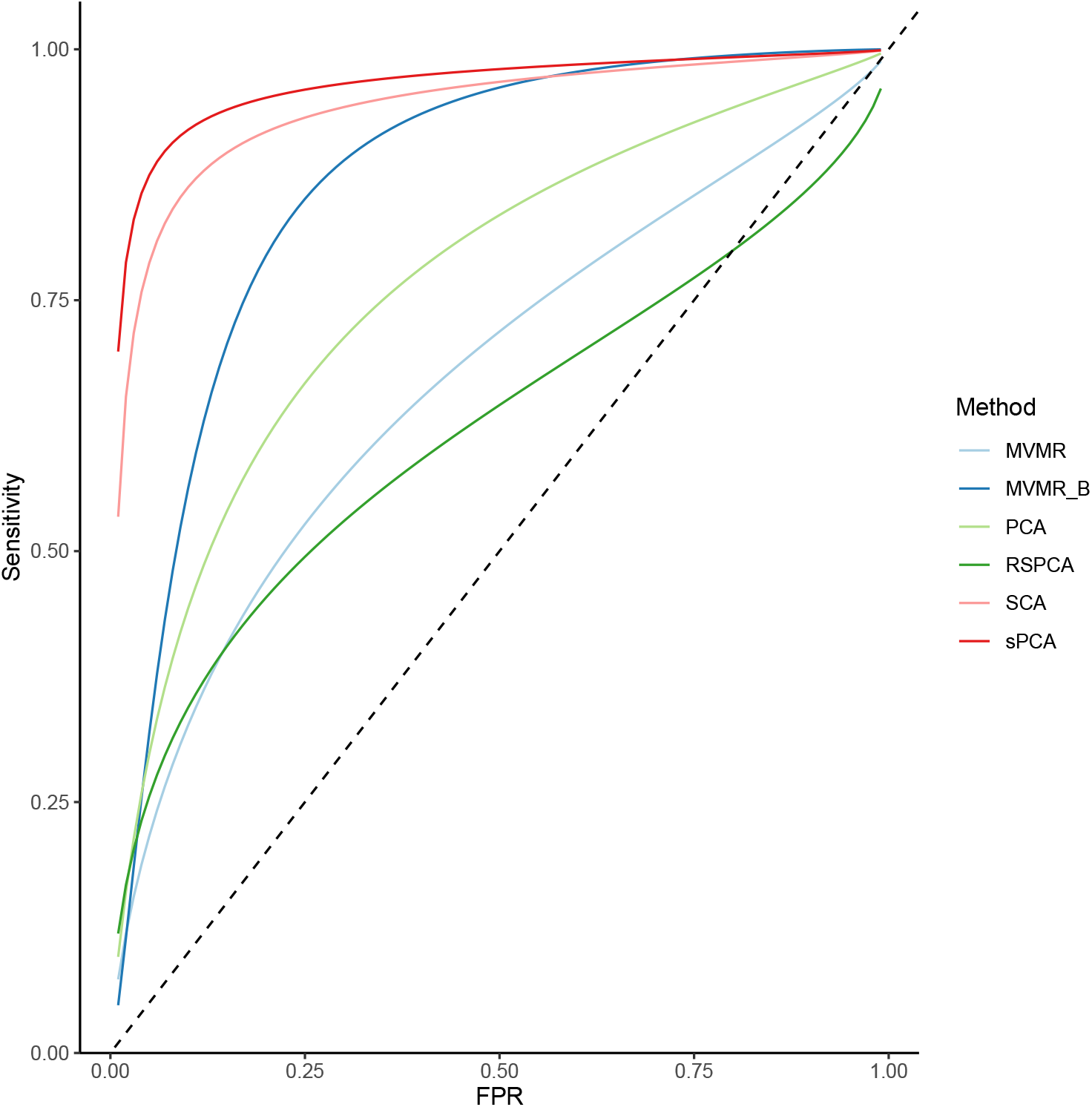
Extrapolated ROC curves for all methods. SCA: Sparse Component Analysis^8^; sPCA: sparse PCA (Zou et al.)^16^; RSPCA: robust sparse PCA^26^; PCA: principal component analysis; MVMR: multivariable MR; MVMR_B: MVMR with Bonferroni correction.

## Results

### Univariable MR (UVMR) & Multivariable MR (MVMR)

A total of 67 traits were associated with CHD at or below the Bonferroni-corrected level (*p* = 0.05*/*118, Table 2). Two genetically-predicted lipid exposures (M.HDL.C, M.HDL.CE) were negatively associated with CHD and 65 were positively associated (Table 1). In a MVMR model including only the 67 Bonferroni-significant traits, fitted with the purpose of illustrating the instability of IVW-MVMR in conditions of high collinearity, the conditional *F*-statistic (*CFS*)^4^ was lower than 2.2 for all exposures (with a mean *CFS* of 0.81), highlighting the weak instrument problem. The causal estimates for two traits (ApoB, M.LDL.PL) reached significance (OR (95% CI): 1.031(1.012, 1.37) and 0.904(0.881, 0.923) (Fig. 9). For M.LDL.PL, its UVMR estimate (1.52(1.35, 1.71), p < 10^−10^)) was of opposite sign to the MVMR estimate. Since these traits are correlated (*r*^2^ = 0.89), we interpret this change of effect direction as an artefact of multicollinearity. Additionally, the ability to detect any further causal effects is likely masked by variance inflation. To see if the application of a weak-instrument robust MVMR method could improve the analysis, we applied MR GRAPPLE^5^). Although the method did not identify any of the exposures as significant at Bonferroni-adjusted significance level, the estimate for M.LDL.PL is still negative but closer to zero and not statistically significant. The only trait that retains statistical significance is ApoB.

**Table 1.**
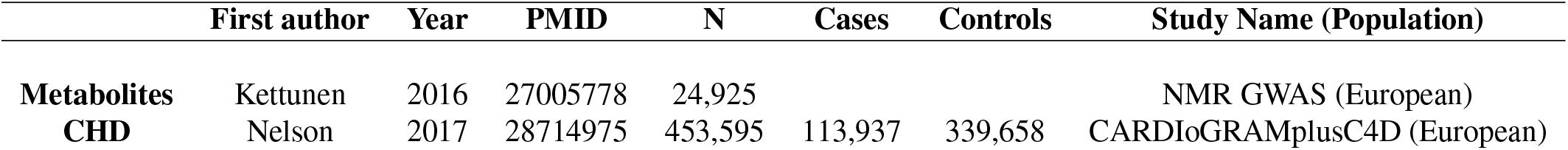
Two-sample MR. Study characteristics.

**Table 2.** Univariable MR results for the Kettunen dataset with CHD as the outcome. Positive: positive causal effect on CHD risk; Negative: negative causal effect on CHD risk.

|  | Positive | Negative |
| --- | --- | --- |
| VLDL | M.VLDL.C, M.VLDL.CE, M.VLDL.FC, M.VLDL.L,<br>M.VLDL.P, M.VLDL.PL, M.VLDL.TG, XL.VLDL.L,<br>XL.VLDL.PL, XL.VLDL.TG, XS.VLDL.L, XS.VLDL.P, XS.VLDL.PL,<br>XS.VLDL.TG, XXL.VLDL.L, XXL.VLDL.PL,<br>L.VLDL.C, L.VLDL.CE, L.VLDL.FC, L.VLDL.L, L.VLDL.P,<br>L.VLDL.PL, L.VLDL.TG, S.VLDL.C, S.VLDL.FC,<br>S.VLDL.L, S.VLDL.P, S.VLDL.PL, S.VLDL.TG | None |
| LDL | LDL.C, L.LDL.C, L.LDL.CE, L.LDL.FC, L.LDL.L, L.LDL.P, L.LDL.PL,<br>M.LDL.C, M.LDL.CE, M.LDL.L, M.LDL.P,<br>M.LDL.PL, S.LDL.C, S.LDL.L, S.LDL.P | None |
| HDL | S.HDL.TG, XL.HDL.TG | M.HDL.C, M.HDL.CE |

### PCA

Standard PCA with no sparsity constraints was used as a benchmark. PCA estimates a square loadings matrix of coefficients with dimension equal to the number of exposures *K*. The coefficients in the first column define the linear combination of exposures with the largest variability (PC1). Column 2 defines PC2, the linear combination of exposures with the largest variability that is also independent of PC1, and so on. This way, the resulting factors seek to reduce redundant information and project highly correlated SNP-exposure associations to the same PC.

In PC1, VLDL-and LDL-related traits were the major contributors (Figure 5a). ApoB received the 8th largest loading (0.1343, maximum was 0.137 for cholesterol content in small VLDL) and LDL.C received the 49th largest (0.1122). In PC2, HDL-related traits were predominant. The first 18 largest positive loadings are HDL-related and 12 describe either large or extra-large HDL traits. PC3 receives its scores mainly from VLDL traits.

Seven components were deemed significant through the permutation-based approach (Fig. 1, *Methods*). This method randomly permutes *γ* and performs the PCA decomposition on each rearranged dataset. Eigenvalues are calculated for each PC based on the permuted matrix. The PCs with an eigenvalue higher than the mean of the permuted sets are deemed informative/ non-trivial and are retained for further analyses. In the second-step IVW regression (Fig. 1), a modest yet significant (OR = 1.002(1.0014, 1.0024), *p <* 10^−10^) association of PC1 with CHD was observed. Conversely, PC3 was marginally significantly associated with CHD (OR = 0.998 (0.998, 0.999), p = 0.049). Since *γ* has been transformed with linear coefficients (visualized in loadings matrix, Fig. 5), estimation of the magnitude of an association is not straightforward; however, significance and orientation of effects can be interpreted. Specifically, when positive loadings are applied to a block of correlated exposures that positively associate with the outcome, the MR estimate is positive; conversely, if negative loadings are applied, the MR estimate is negative.

### Sparse PCA methods

We next employed multiple sparse PCA methods (Table 3) that each shrink a proportion of loadings to zero. The way this is achieved differs in each method. Their underlying assumptions and details on differences in optimisation are presented in Table 3 and further described in Methods.

**Table 3.** Overview of sPCA methods used. KSS: Karlis-Saporta-Spinaki criterion.

| Package | Authors | Features | Choice | PCs |
| --- | --- | --- | --- | --- |
| *elasticnet* | Hui Zou, Trevor Hastie<sup>16</sup> | Formulation of sPCA as a regression problem | KSS | 7 |
| *pcaPP* | Peter Filzmoser, Heinrich Fritz, Klaudius Kalcher<sup>26</sup> | Robust sPCA (RSPCA) with a different measure of dispersion ($Q_n$) | Permutation KSS | 7 |
| Sparse Fused PCA | Jian Guo, Gareth James, Elizaveta Levina, George Michailidis, Gi Zhu | Fused penalties for block correlation | KSS | 14 |
| SCA | Fan Chen, Karl Rohe<sup>8</sup> | Rotation of eigenvectors for approximate sparsity | Permutation KSS | 7 |

The loadings for all methods (Figures 5a-e) are presented for LDL.c, a previously prioritised trait^27^. We do not expect the loadings for the same component to be numerically identical across methods, eg. an HDL-dominant component may be ranking second (in variance explained) in one method and fourth in another. Ideally, we would like to see metabolites contributing to a a small number or only one component. Using a visualisation technique proposed by Kim et al.^28^, this is indeed observed in Fig. 12. This visualisation was also repeated for urea, a metabolite that was assumed to be of little contribution in the overall variance of the (mainly lipid trait-focused) NMR dataset. Urea receives loadings of exactly zero or near zero in all methods (Fig. 12), thus serving as a successful negative control. In the second-step IVW meta-analysis, it appears that the PCs that comprise of predominantly VLDL/LDL and HDL traits robustly associate with CHD, with differences among methods (Table 4).

**Table 4.** Results for dimensionality reduction approaches. Overlap: Percentage of metabolites receiving non-zero loadings in ≥ 1 component. Overlap in PC1, PC2: overlap as above but exclusively for the first two components which by definition explain the largest proportion of variance. VLDL, LDL and HDL significance: results of the IVW regression model with CHD as the outcome for the respective sPC’s (the sPC’s that mostly received loadings from these groups). The terms VLDL and LDL refer to the respective transformed blocks of correlated exposures; for instance, VLDL refers to the weighted sum of the correlated VLDL-related *γ* associations, such as VLDL phospholipid content and VLDL triglyceride content. †: RSPCA projected VLDL-and LDL-related traits to the same PC (sPC1). ‡: SCA discriminated HDL molecules in 2 sPC’s, one for traits of small-and medium-sized molecules and one for large-and extra-large-sized. †: significance is not directly reported in this model.

|  | PCA | sPCA Zou | RSPCA | SFPCA | SCA |
| --- | --- | --- | --- | --- | --- |
| Overlap | 100 | 52.54 | 74.58 | 100.00 | 28.81 |
| Overlap in PC1, PC2 | 100 | 80.51 | 78.81 | 97.46 | 46.61 |
| % Sparse | 0 | 25.79 | 4.72 | 0.18 | 20.94 |
| VLDL, LDL significance in MR | Yes | Yes | Yes | No† | Yes |
| HDL significance in MR | Yes | No | No | No | Yes‡ |

**Figure 9.**
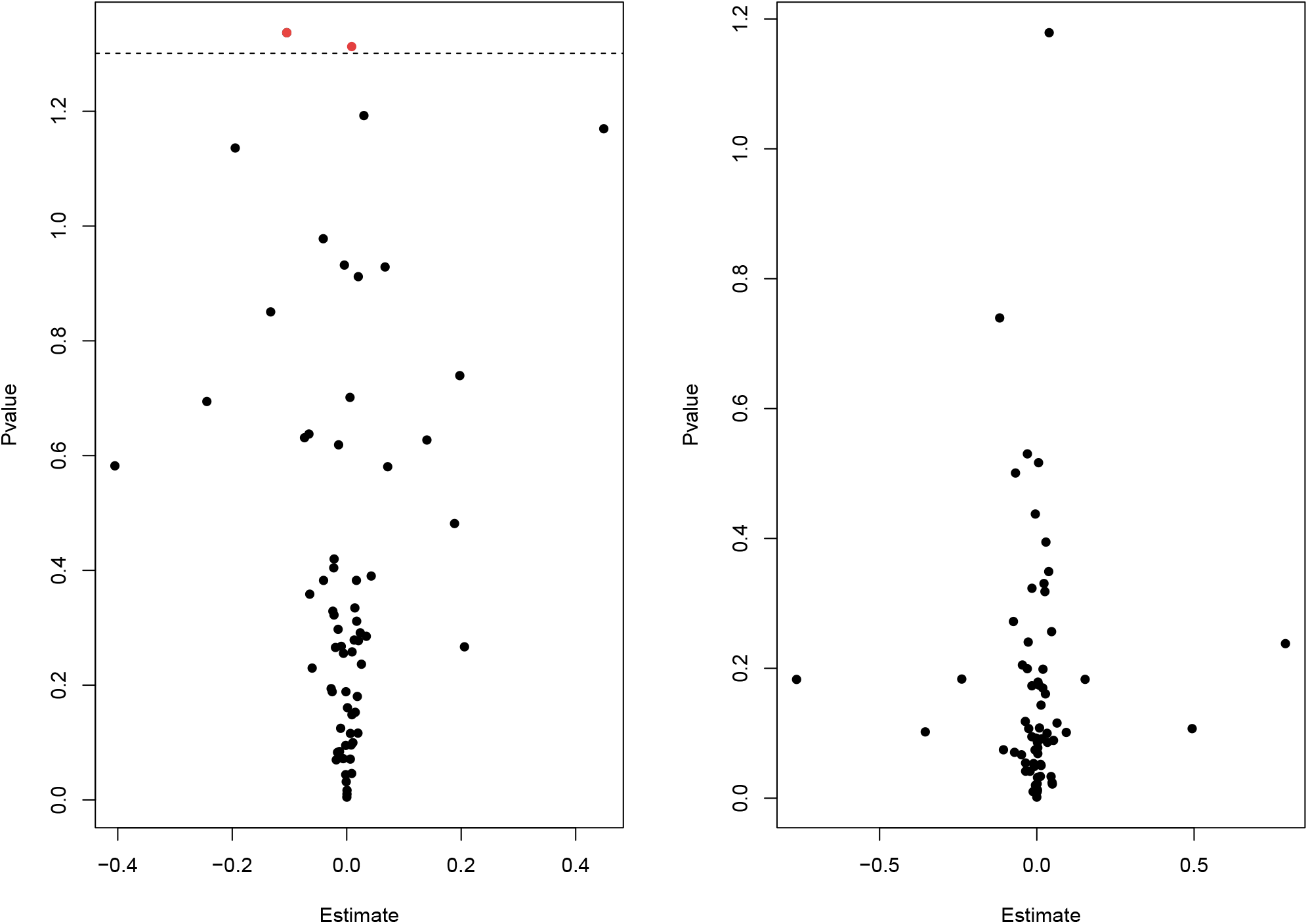
MVMR with IVW (left) and MVMR with GRAPPLE^33^ (right). Only the 67 exposures that are significant in UVMR are put forward in these models. In IVW (left), M.LDL.PL shows nominal significance. In MR GRAPPLE (right), apolipoprotein B has the lowest p-value but no trait reaches nominal significance.

**Figure 10.**
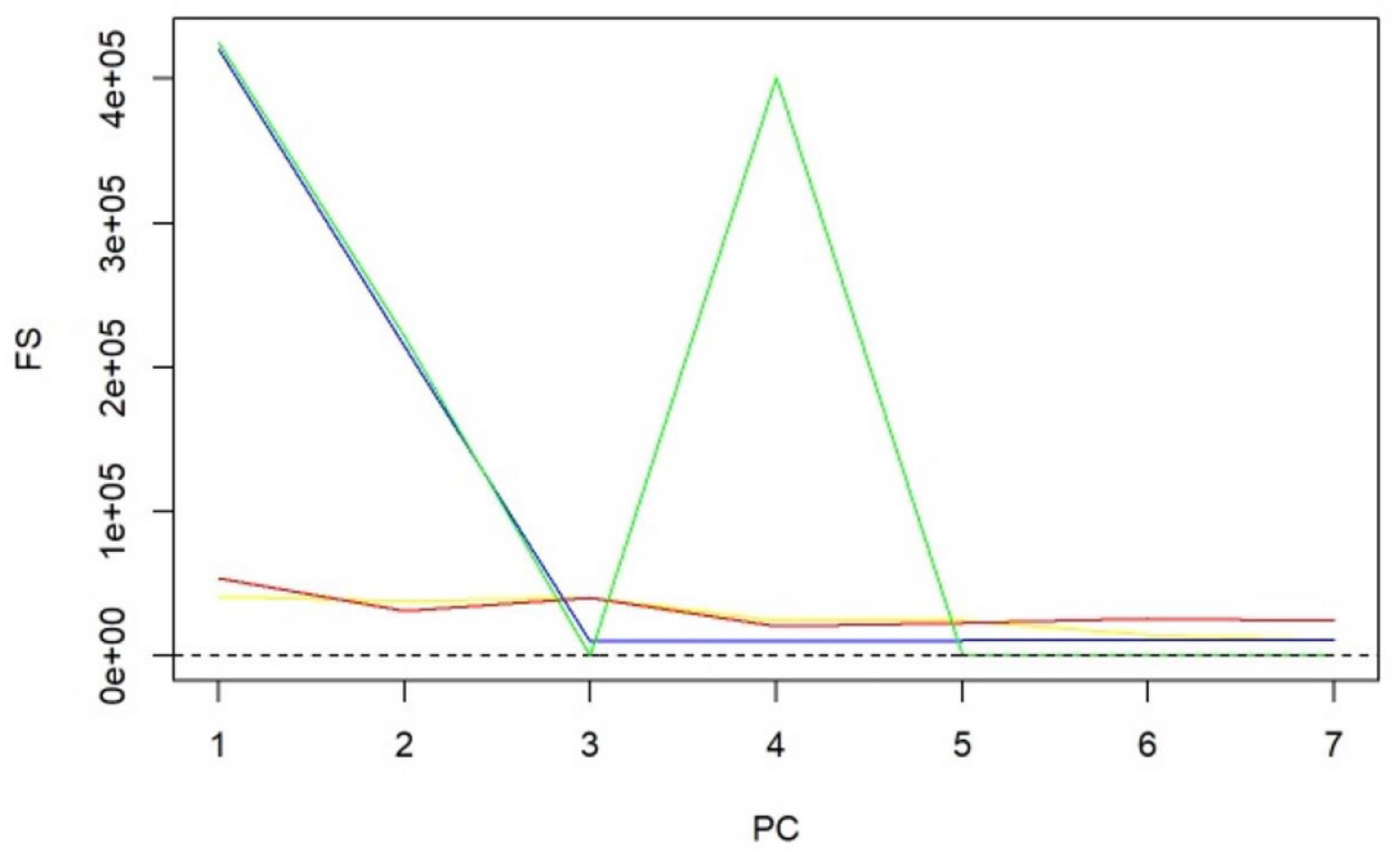
*F*-statistics for PCs and sparse PCs. The formula derived in Eq. 4 is used. Black: PCA (no sparsity constraints); Yellow: SCA; Red: sparse PCA (Zou); Blue: Sparse robust PCA; Green: Sparse fused PCA. The dashed line represents the cutoff of 10 that is considered the minimum desired *F*-statistic for an exposure to be considered well instrumented. The green line diverges from the pattern of decreasing instrument strength but, when referring to the loadings heatmap (Figure 5), it can be observed that the 4th sparse PC in the fused sPCA receives negative loadings from multiple VLDL and LDL related traits. This may in turn cause the large *F*-statistic.

**Figure 11.**
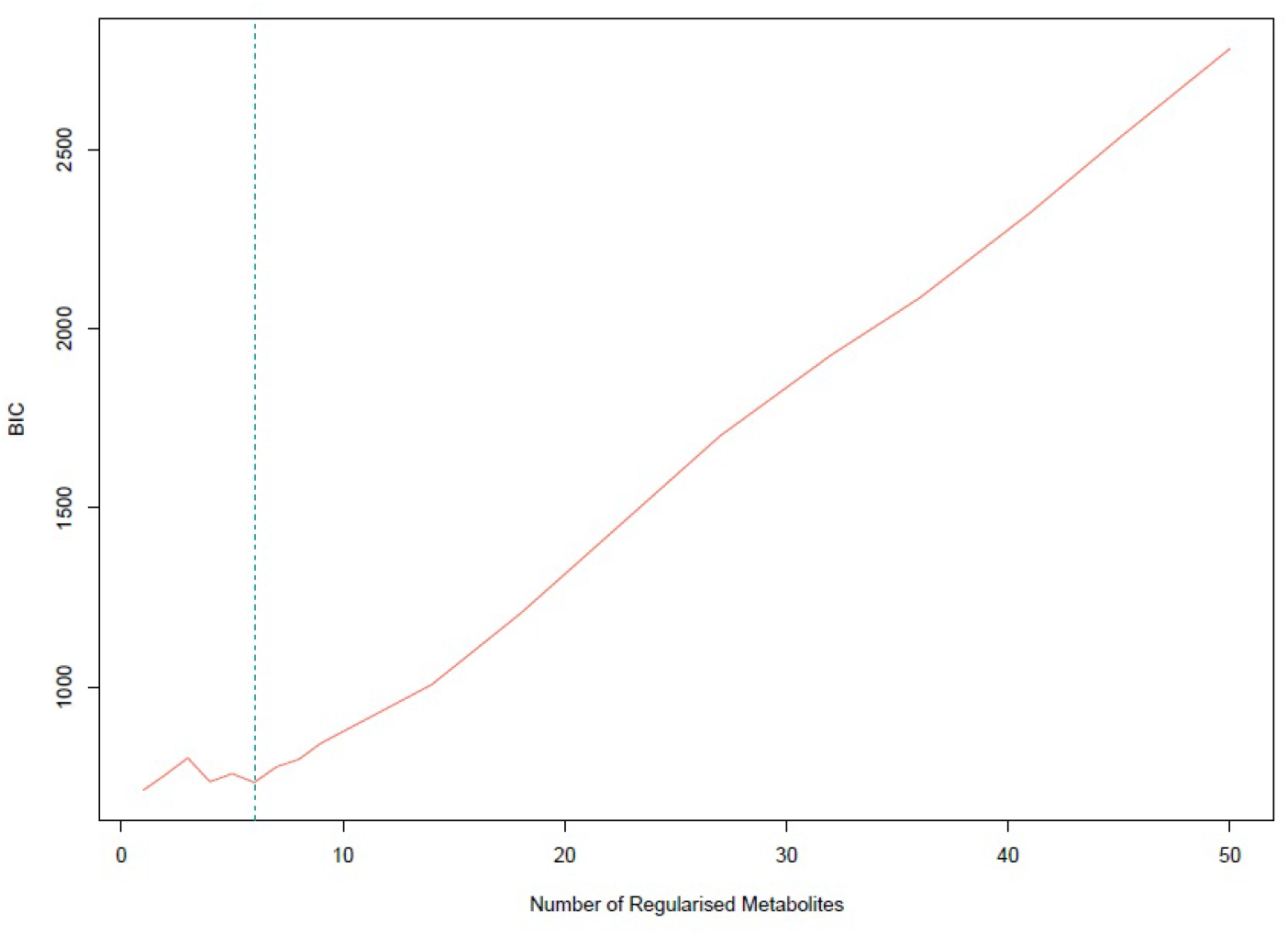
: Bayesian Information Criterion (BIC) for different numbers of metabolites regularized to 0. The lowest value is achieved for one non-zero exposure per component. However, six non-zero exposures per component also achieved a similar low BIC and this was selected.

**Figure 12.**
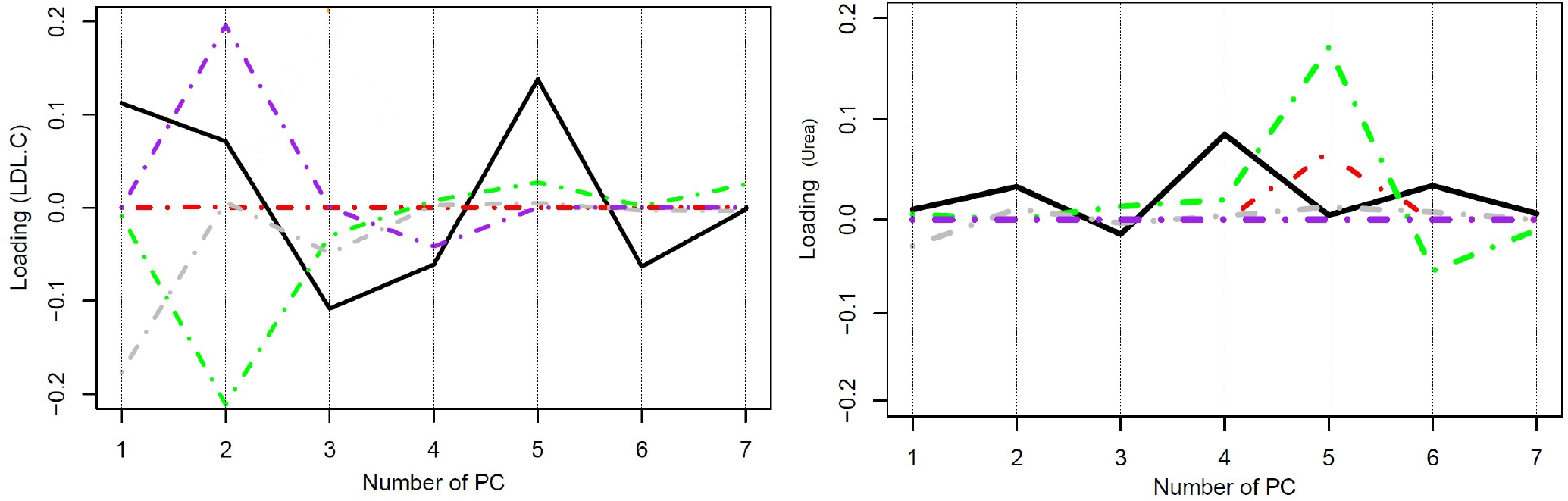
Trajectories for the loadings of total cholesterol in LDL and urea in all methods. In LDL.C, PCA. In the urea plot, in SCA and sPCA this metabolite is regularised to 0 throughout all sPC’s. It appears that the RSPCA loadings (green) are the only ones larger in magnitude than in unconstrained PCA, although in the first 2 sPC’s they are equal to 0. Black solid: Unconstrained PCA, red: sPCA (Zou et al., 2006), green: RSPCA, grey: sparse fused PCA, purple: SCA.

### Sparse PCA (Zou et al., 2006)

This method adapts the PCA decomposition to a ridge regression and adds an *L*_1_ norm to introduce sparsity^16^. In order to get an estimate for the degree of sparsity to be induced, a grid search was performed. Multiple sPCA models with different numbers of sparse traits per component were generated and the lowest Bayesian Information Criterion (BIC) was achieved for a total of six metabolites not regularised to 0 per each sPC. While the minimum BIC is achieved when one metabolite is not regularised to 0, 6 non-zero loadings were preferred as this appears to be an inflection point (Figure 11). Under this level of sparsity, the permutation-based approach suggests that seven sPC’s are to be retained. Seventy-one exposures received a zero loading across all components. The first sPC is comprised of VLDL traits, ApoB and triglycerides (Fig. 5b) and is estimated to exert a significant causal effect on CHD (OR (95% CI): 1.003(1.0003, 1.005)). The sPCs 2 to 7 were not associated with CHD (range: 0.999(0.998, 1.001), 1.003(1.000, 1.062), all *p* values > 0.05).

### RSPCA

This approach is due to Croux et al.^26^. Apart from sparsity, the authors also use an alternative measure of dispersion, the *Q*_*n*_ estimator^18^, that is robust to outliers in *γ*. Optimisation and the Karlis-Saporta-Spinakis (KSS) criterion pick seven components to be informative^9^. KSS is a less computationally expensive method (compared to the permutation method) for defining an eigenvalue threshold for PC inclusion^9^. The overlap of the PCs is visualised in Figure 5c. Large positive loadings are observed for VLDL-and LDL-related traits in sPC1 and five HDL-related traits are regularised to 0. LDL.C and ApoB received the 9th and 12th largest positive loadings. In the second sPC, HDL traits are mainly represented. None of the sPCs are robustly associated with CHD.

### Sparse Fused PCA (SFPCA)

SFPCA works by assigning highly correlated variables the exact same loadings as opposed to numerically similar ones (Figure 5d). This is achieved with two norms in the objective function: *λ*_1_ which regulates the *L*_1_ norm that induces sparsity and *λ*_2_ for the *L*2 regularisation (squared magnitude of *γ*) to guard against singular solutions. A grid search is used to identify appropriate parameters for *λ*_1_ and *λ*_2_. Applying the KSS criterion indicated that 6 sPCs should be retained. The loadings matrix is presented in Figure 5d and the identical coloring visualises the unique loadings in sPCs 1, 3 and 4, with only one metabolite (M.HDL.P) received a zero loading in sPC 1. Two large clusters are apparent; one with HDL-related molecules and one with VLDL-, LDL-, IDL-related and other molecules. Both ApoB and LDL.C received the largest negative loading (−0.11). This was also received by 43 other metabolites (26 VLDL-related, 9 LDL-related, 6 IDL related, Serum TG, S.HDL.TG). The first 2 sPC’s were significantly negatively associated with CHD (0.998(0.997, 0.999), *p* value = 6.63*x*10^−7^ and 0.9989(0.9980, 9998), *p* value = 0.016). Our results show that we generally retain the expected direction of effects by examining the factor loadings. Here, the component that consists of the weighted sum of VLDL and LDL related traits (sPC1) is *negatively* associated with CHD because of the negative loadings (Fig. 5d, in red).

### Sparse Component Analysis (SCA)

SCA performs an initial rotation of the obtained loadings in order to achieve approximate sparsity^8^. Seven components were retained after a permutation test. In the final model, more than 60 metabolites were regularised to zero in all components. In the heatmap, little overlap is noted among the metabolites (Figure 5e). The first sPC mainly receives loadings from LDL and IDL, sPC2 from VLDL and sPC3 from large and extra-large HDL traits. sPC6 gathers the largest loadings for LDL, the fifth for VLDL and the contribution of HDL is split in sPC’s 3 (large and extra-large HDL traits) and 4 (small and medium HDL traits). sPC1 was positively associated with CHD (OR (95% CI), *p* value:1.013(1.009, 1.017), *<* 10^−8^). sPC4, receiving loadings mainly from medium-sized HDL molecules, was negatively associated with CHD (0.997(0.995, 0.999), 0.02), while sPC3 (large and extra-large HDL traits), was positively associated.

### Comparison with Univariable MR

In principle, all PC methods derive independent components. This is strictly the case in PCA, where subsequent PCs are orthogonal, and is relaxed to some degree in sparse implementations. We hypothesised that UVMR and MVMR could provide similar causal estimates of the associations of metabolite PCs with CHD. The results are presented in Figure 6 and concordance between UVMR and MVMR is quantified with the *R*^2^. The largest agreement of the causal estimates is observed in PCA. In the sparse methods, SCA^8^ and sPCA^16^ provide similarly consistent estimtates, whereas some disagreement is observed in the estimates of PC4 and

### Instrument Strength

Instrument strength for the chosen PCs was assessed via an *F*-statistic, calculated using a bespoke formula that accounts for the PC process (see Methods and Appendix). The *F*-statistics for all transformed exposures cross the cutoff of 10. There was a trend for the first components being more strongly instrumented in all methods (see Figure 10). In the individual exposures, the conditional *F*-statistic for all exposures is lower than three for all exposures. Thus it appears that, while individual exposures are weakly jointly instrumented with the current instrument, PCs in contrast exhibit an improved instrument strength.

### Simulation Study

We investigated the efficiency and reliability of various PCA methods compared to standard MVMR and Bonferroni-corrected MVMR in a simulation setting. *K* = 30−60 exposures within *b* = 4−6 blocks were generated. Exposures within blocks were genetically correlated and all exposures were correlated due to shared environmental confounding. A simplified causal diagram consistent with the data generating mechanism is presented in Fig. 7 for the case of only *K* = 6 exposures and *b* = 2 blocks. Only some of the exposures exert a true causal effect on the outcome. (e.g. *X* 1 and *X* 2 in Fig. 7).

In order to compare the results of a PC regression with the results from an MVMR with all exposures, we interpret the dimensionality reduction methods as tests. We then define true positive, true negative, false positive and false negative results. For the MVMR method, a true positive (*TP*) is an exposure that is causal in the underlying model (i.e *X*_1_ and *X*_2_ in Fig. 7) *and* is identified as statistically significant by MVMR. Likewise, a true negative (TN) would be a non causal exposure (e.g. *X*_4_) that is not statistically significant in the MVMR model.

In the PCA and sparse PCA methods, this classification is determined with respect to the causal estimate of a specific PC. If a PC is determined by the correlated exposures *X*_1_, *X*_2_ and *X*_3_ *and* it has a statistically significant causal estimate, then *X*_1_ and *X*_2_ are true positives and *X*_3_ is false positive. Similarly, if the PC constructed by *X*_4_, *X*_5_ and *X*_6_ is not significant in an MVMR analysis, all three would count as true negatives as they do not causally affect *Y*.

Across a range of simulation scenarios we collated summary data on *TP, FP, TN* and *FN* rates for each method, by averaging over the individual results for each exposure. This included varying the sample sizes (*n* = 1000−40000, which in turn corresponded to a mean instrument strength in *γ* of between 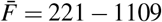 and mean conditional F statistic 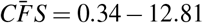. We then derived the sensitivity (SNS), specificity (SPC) and false positive rate (FPR) as

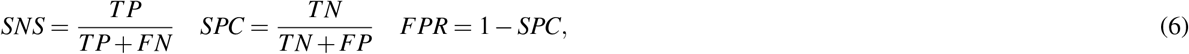

and compared all methods by their area under the receiver-operating characteristic (ROC) curve (*AUC*) and the Youden’s index *J* (*J* = *SNS* + *SPC*−1). Specifically, the *AUC* was derived from the extrapolated *ROC* curve using the method of Reitsma et al.^29^, which jointly meta-analyses *SNS* and *SPC*.

Two sparse PCA methods (SCA^8^, sPCA^16^) consistently achieve the highest AUC (Fig. 8). This advantage is mainly driven by an increase in sensitivity for both these methods compared with MVMR. A closer look at the individual simulation results corroborates the discriminatory ability of these two methods, as they consistently achieve high sensitivities and most 14. Both standard and Bonferroni-corrected MVMR performed poorly in terms of AUC (AUC 0.660 and 0.885 respectively), due to poor sensitivity. While there was a modest improvement with PCA (AUC 0.755), it appears that PCA and RSPCA do not accurately identify negative results (median specificity 0 and 0.28 respectively). This extreme result can be understood by looking at the individual simulation results in Figure 14; both PCA and RSPCA cluster to the upper right end of the plot, suggesting a consistently low performance in identifying true negative exposures. In specific, the estimates with both these methods were very precise across simulations and this resulted in many false positive results and low specificity. We note a differing performance among the top ranking methods (SCA, sPCA); while both methods are on average similar, the results of SCA are more variable in both sensitivity and specificity (Table 5). The Youden indexes for these methods are also the highest (Fig. 8a).

**Figure 13.**
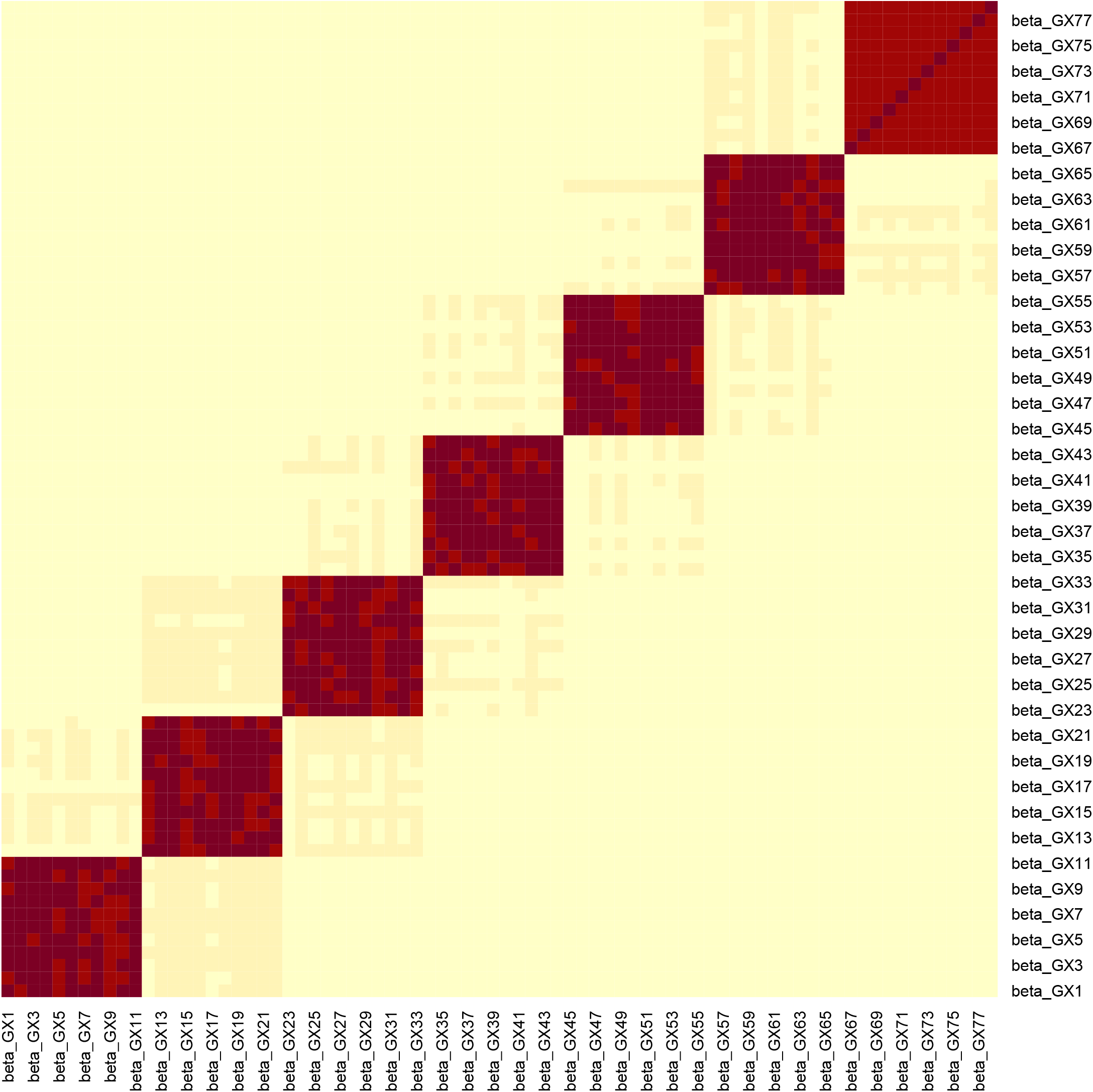
Example for the block correlation in *γ* (*n* = 5000, *K* = 77) induced by the data generating mechanism in Fig. 7. In this example, the mean *F*-statistic is 231.2 and the mean *CFS* is 3.21.

**Figure 14.**
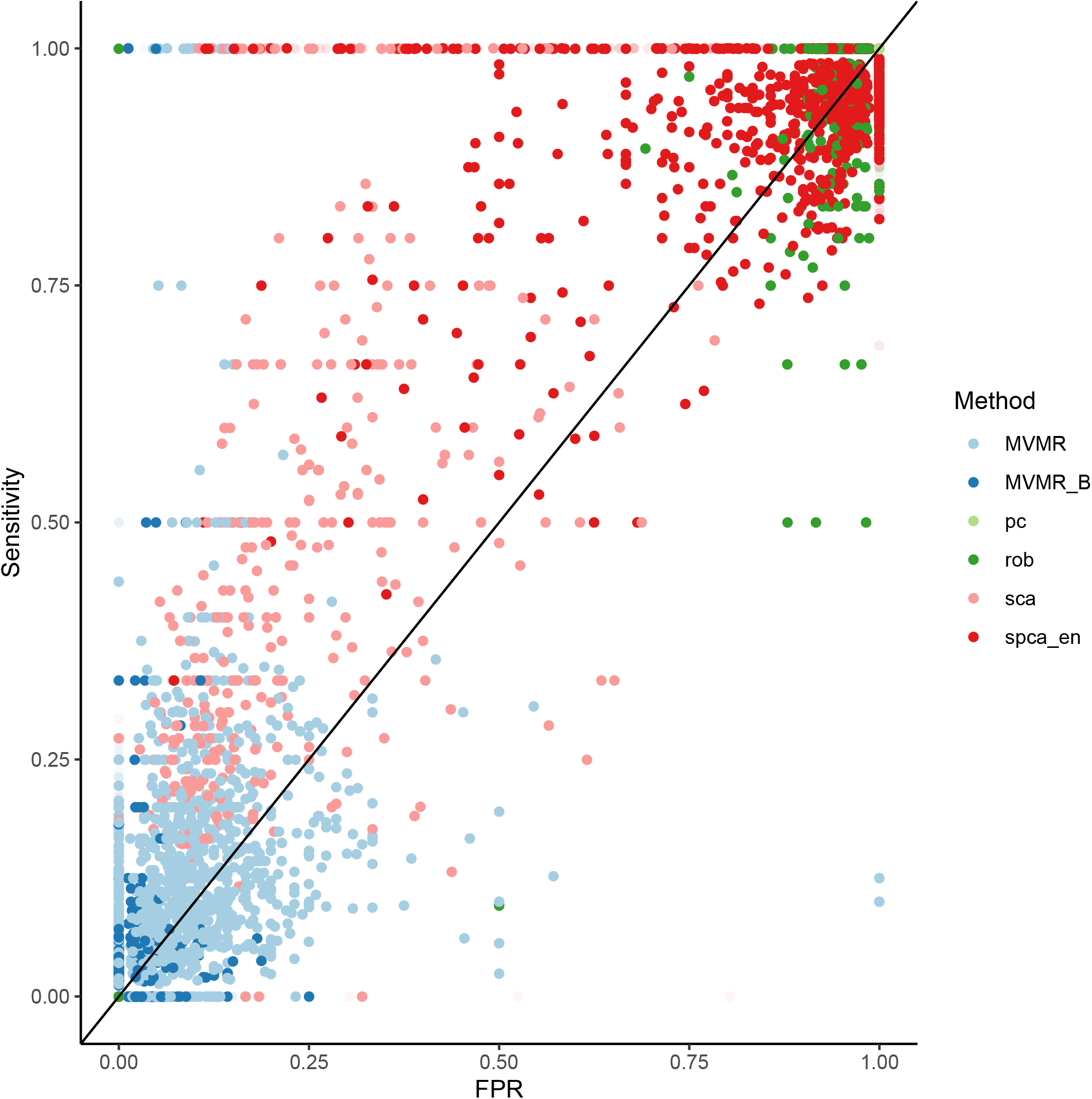
Individual Results from *s* = 1000 simulations.

**Table 5.** Sensitivity & Specificity presented as median and interquartile range across all simulations. Presented as median sensitivity/specificity and interquartile range across all simulations; *AUC*: area under the ROC curve.

|  | PCA | SCA | sPCA | RSPCA | MVMR | MVMR_B |
| --- | --- | --- | --- | --- | --- | --- |
| **AUC** | 0.755 | 0.925 | 0.947 | 0.611 | 0.660 | 0.854 |
| **Sensitivity** | 1,0.1 | 1,0.21 | 1, 0.047 | 0.667, 0.251 | 0.222, 0.2 | 0, 0.076 |
| **Specificity** | 0,0.02 | 0.925,0.772 | 0.925, 0.041 | 0.278, 0.111 | 0.960, 0.048 | 1,0 |
| **Youden's J** | 0 | 0.584 | 0.778 | -0.061 | 0.192 | 0.044 |

Even with large sample sizes (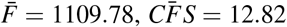 for a sample size *n* = 6000), MVMR can still not discriminate between positive and negative exposures as robustly as the sparse PCA methods. A major determinant of the accuracy of these methods appears to be the number of truly causal exposures, as in a repeat simulations with only four of the exposures being causal there was a drop in sensitivity and specificity across all methods. Sparse PCA methods still outperformed other methods in this case, however (Table 6).

**Table 6.** Simulation study on only four exposures (out of the total *K* = 50) contributing to the outcome *Y*. A drop in sensitivity and specificity is observed for SCA and sPCA compared with the simulation configuration in Table 5.

|  | PCA | SCA | sPCA | RSPCA | MVMR | MVMR_B |
| --- | --- | --- | --- | --- | --- | --- |
| **AUC** | 0.863 | 0.714 | 0.859 | 0.492 | 0.511 | 0.675 |
| **SNS** | 1,0.03 | 0.75,0.25 | 1,0.17 | 0.5,0.25 | 0.25,0.25 | 0,0 |
| **SPC** | 0,0.2 | 0.76,0.46 | 0.66,0.18 | 0.37,0.15 | 0.94,0.07 | 1,0 |
| **Youdens J** | 0 | 0.428 | 0.625 | -0.029 | 0.105 | 0.032 |

## Discussion

We propose the use of sparse PCA methods in MVMR in order to reduce high-dimensional exposure data to a low number of PCs and infer the latter’s causal contribution. As the dimensionality of available data sets for MR investigations increases (e.g. in NMR experiments^30^ and imaging studies), such approaches are becoming ever more useful. Our results support the notion that sparse PCA methods retain the information of the initial exposures. Specifically, the SCA^8^ and sPCA^16^ methods, performed best in simulation studies, whereas the SCA approach performed best in the positive control example of lipids and CHD.

By employing sparse PCA methods in a real dataset^6^, we show that the resulting PCs group VLDL, LDL and HDL traits together, whilst metabolites acting via alternative pathways receive zero loadings. This is a desirable property and indicates that the second-step MR enacted on the PCs obtains causal estimates for intervening on biologically meaningful pathways.^31^. This is in contrast with unconstrained PCA, in which all metabolites contribute to all PCs. Previously, Sulc et al. used PCA in MR to summarise highly correlated anthropometric variables^32^. To our knowledge, this is the first investigation of different sparse PCA modalities in the context of MR. We additionally provide a number of ways to choose the number of components in a data-driven manner. Our proposed approach of a sparse PCA method could potentially reduce overlap across components; for instance, in the paper by Sulc et al., there are PCs that are driven in common by trunk, arm and leg lean mass, basal metabolic rate and BMI. This is an important topic for further research.

This approach is conceptually different from the robust methods that have been developed for standard multivariable MR in the presence of weak instruments, such as MR GRAPPLE^5^. Our method reduces the need for a pre-selection of which exposures to include in a MVMR model. We present a complementary workflow through which we can include all available exposures with no prior selection, collate them in uncorrelated and interpretable components and then investigate the causal contribution of these groups of exposures and avoids the risk of generating spurious results in such an extreme setting of high collinearity compared with MVMR IVW and MR GRAPPLE formulations. For example, a 2019 three-sample MR study that assessed 82 lipoprotein subfraction risk factors’ effects on CHD used a UVMR and a robust extension of MVMR. A positive effect of VLDL-and LDL-related subfractions on CHD was reported, consistent in magnitude across the sizes of the subfractions^33^. Results were less definitive on the effect of HDL subfractions of varying size on CHD. Both positive and negative effects were observed. In our study, the HDL subfractions were uniformly projected to similar subspaces, yielding a single component that was mainly HDL populated in all models, except for the SCA model 15 which projected the small/ medium and large/ extra-large HDL traits in two different components. In all cases, the association of the sPCs with CHD was very low in magnitude. Nevertheless, the direction of effects was in line with the established knowledge on the relationship between lipid classes and CHD.

In the sparse PCA methods, there were differences in the results. The sPCA method^16^ favoured a sparser model in which less than 10 metabolites per PC were used. This observation is also made by Guo et al^21^. The SCA method^8^ achieved good separation of the traits and very little overlap was observed. A separation of HDL-related traits according to size, not captured by the other methods, was noted. Clinical relevance of a more high-resolution HDL profiling, with larger HDL molecules mainly associated with worse outcomes, has been previously reported^34^.

### Limitations

In the present study, many tuning parameters needed to be set in order to calibrate the PCA methods. We therefore caution against extending our conclusions on the best method outside the confines of our simulation. Not all available sparse dimensionality reduction approaches were assessed in our investigation and other techniques could have provided better results.

The sparse PCA approach outlined in this paper enables the user to perform a MVMR analysis with a large number of highly correlated exposures, but one downside is that the effect sizes are not as interpretable. Of course, if a proven pharmaceutical or lifestyle intervention exists which simultaneously affects a number of lipid fractions, and those fractions are also combined within a single principle component, then arguably the PC approach’s causal estimate is more relevant than that of a single lipid fraction in a multivariable model. Further foundational work on lipid metabolism pathways could shed light on this.

### Conclusion

In the present study, we underline the utility of sparse dimensionality reduction approaches for highly correlated exposures in MR. We present a comprehensive review of methods available to perform dimensionality reduction and describe their performance in a real data application and a simulation.

## Data Availability

This is a computational study and no new data were generated. The code for the analyses is publicly available.

https://github.com/vaskarageorg/SCA_MR

## Data Availability

The GWAS summary statistics for the metabolites (http://www.computationalmedicine.fi/data/NMR_GWAS/) and CHD (http://www.cardiogramplusc4d.org/) are publicly available. We provide code for the SCA function, the simulation study and related documentation on github (https://github.com/vaskarageorg/SCA_MR/).

## Appendix

### MVMR with IVW and GRAPPLE

A small negative effect for M.LDL.PL is noted as nominally significant in Fig. 9. This is not concordant with the UVMR direction of effect. In GRAPPLE, no traits surpass the nominal significance threshold.

### Simulation Study Design

We generate a minor allele frequency *MAF* ∼*U* (0.01, 0.5) and an unmeasured confounder *U* ∼*N*(0, 5). Conditionally on the MAF, we generate the genes *G* ∼*Bin*(2, *MAF*). We define *Q* blocks and divide the *P* genes of *G* to *q* blocks, each containing *P/Q* genes. This approach aims to model the biological phenomenon of multiple traits being influenced by a common set of variants. For instance, the traits of LDL cholesterol content and LDL phospholipid content appear to be associated with the variants in a largely similar manner^6^.

For each block *q* = 1, 2, …, *Q*, we define a matrix *γ*_*q*_ whose elements *γ*_1_−*γ*_*P/Q*_ are non-zero and ∼*N*(4, 1). This matrix is what will direct only certain variants to influence certain exposures, leading to multicollinearity. We then generate the *K* exposures sequentially per block, following the parametric equation *X*_*q*_ = *γ*_*q*_*G*_*q*_ + *U* + *ε*_*X*_. This way, specific genetic variants generate blocks of exposures as shown in Fig. 7 and the exposures *X*_1_ −*X*_*K/Q*_ are highly correlated. We derive the SNP-exposure associations from this dataset as 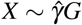. We set *K*=50-100 and *P*=100-150 and the blocks 5-8. We let these values vary across simulations in order to generate more varying values in diagnostic accuracy. For a given sample size, *s* = 1000 simulation studies were performed.

To retain the workflow of a two-sample MR, we generate a second exposure set identically as above but independent. This step is important as it guarantees the no sample overlap assumption of two-sample MR^35^. Based on this second *X* ^′^ matrix, we generate the outcome *Y* = *β X* ^′^ + *U* + *ε*_*Y*_. The vector of causal estimates *β* is generated based on any number of exposures being causal in the two blocks. This includes the null. We obtain the SNP-outcome associations as 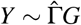.

The effect of the data generating mechanism in a single dataset is visualised in Fig. 13. The methods employed are

- MVMR IVW with 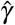 and 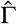^36^.
- MVMR IVW with Bonferroni correction.
- PCA with the scores from 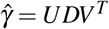.
- sparse PCA as implemented in the *elasticnet* package^16^.
- RSPCA^26^.
- SCA^8^.

Due to computational complexity, sparse PCA in the *elasticnet* package (spca_en in Fig. 8) was implemented with a simplification regarding the sparsity parameter. In specific, it was assumed that the number of non-zero exposures per component was *P/Q*. For SCA, we use the cross-validation method in *PMA R* package^37^.

To generate the summary ROC curves presented in Fig. 8, we treated simulation results as individual studies and meta-analysed them with the bivariate method of Reitsma et al.^29^ in the *R* package. The logit sensitivity and specificity (which are correlated) are jointly meta analysed by modelling them as a bivariate normal distribution and employing a random-effects model for accomodating this correlation and framing it as heterogeneity.

The results for a set simulations in *N* = 3000 individuals are presented in Figure 14. We observe that MVMR and MVMR with Bonferroni correction estimates cluster to the bottom left of the plot, suggesting a low sensitivity. The estimates from SCA and sPCA_EN are spread in the upper left half of the plot and often achieve a sensitivity of 1.00. The PCA and RSPCA mainly provide highly sensitive estimates but perform relatively worse in specificity.

## Notes

### Competing Interest Statement

Dr D.G. is a part-time employee of Novo Nordisk. The other authors have no relevant conflict of interest to declare.

### Funding Statement

VK and Pr. JB received funding from the Expanding Excellence in England (E3) fund.

